# The Coronavirus 2019 pandemic in Canada: the impact of public health interventions on the course of the outbreak in Alberta and other provinces

**DOI:** 10.1101/2020.05.09.20096636

**Authors:** Mohamed Mahsin, Seungwon Lee, David Vickers, Alexis Guigue, Tyler Williamson, Hude Quan, Robert R. Quinn, Pietro Ravani

## Abstract

Background:

The SARS-CoV-2 disease 2019 (COVID-19) pandemic has spread across the world with varying impact on health systems and outcomes. We assessed how the type and timing of public-health interventions impacted the course of the outbreak in Alberta and the other Canadian provinces.

Methods:

We used publicly-available data to summarize rates of laboratory data and mortality in relation to measures implemented to contain the outbreak and testing strategy. We estimated the transmission potential of SARS-CoV-2 before the state of emergency declaration for each province (R_0_) and at the study end date (R_t_).

Results:

The first cases were confirmed in Ontario (January 25) and British Columbia (January 28). All provinces implemented the same health-policy measures between March 12 and March 30. Alberta had a higher percentage of the population tested (3.8%) and a lower mortality rate (3/100,000) than Ontario (2.6%; 11/100,000) or Quebec (3.1%; 31/100,000). British Columbia tested fewer people (1.7%) and had similar mortality as Alberta. Data on provincial testing strategies were insufficient to inform further analyses. Mortality rates increased with increasing rates of lab-confirmed cases in Ontario and Quebec, but not in Alberta. Ro was similar across all provinces, but varied widely from 2.6 (95% confidence intervals 1.9-3.4) to 6.4 (4.3-8.5), depending on the assumed time interval between onset of symptoms in a primary and a secondary case (serial interval). The outbreak is currently under control in Alberta, British Columbia and Nova Scotia (R_t_ <1).

Interpretation:

COVID-19-related health outcomes varied by province despite rapid implementation of similar health-policy interventions across Canada. Insufficient information about provincial testing strategies and a lack of primary data on serial interval are major limitations of existing data on the Canadian COVID-19 outbreak.

## Introduction

In December 2019, an epidemic of the novel coronavirus SARS-CoV-2, the causative agent of Coronavirus disease (COVID-19), emerged in Wuhan, China.^1^ Compared to other highly pathogenic human coronaviruses (MERS-CoV and SARS-CoV), SARS-CoV-2 has a lower case-fatality rate but spreads more efficiently.^2^ As of May 7, 2020, almost 4 million have been infected and over 250,000 have died globally.^3^ Since the index case arrived in Canada from China on January 22, 2020,^4^ the infection has spread across all Canadian provinces and territories. Over one million Canadians have been tested, 65,000 have tested positive for COVID-19 and 4,500 have died.^4,5^ The Government of Canada projections anticipated 11,000–22,000 deaths over the course of the pandemic, assuming "stronger epidemic control", defined by a high degree of physical distancing, a high proportion of cases identified and isolated, and a high proportion of contacts traced and quarantined.^6^

With COVID-19, Canadian Provincial governments have been enforcing unprecedented quarantines and social distancing measures to contain the infection, by reducing interpersonal contact (travel restrictions, physical distancing and self-isolation) and lowering the probability of virus transmission (hand and respiratory hygiene, use of personal protective equipment and surface cleaning). As compared to other regions, Western Canadian provinces implemented public education initiatives and policy measures relatively early in response to the first identified COVID-19 cases. How these interventions impacted the outbreak in terms of both common measures of infectivity and important health outcomes is unknown.

We studied the transmission potential of SARS-CoV-2 virus before and after the implementation of measures designed to contain the infection in Alberta and compared data to other Canadian provinces.^7^ Testing strategies may have changed in response to the identification of new infection foci, and testing strategies could impact case ascertainment. As a result, we studied the rates of confirmed cases as well as hospital admission and death rates, which are expected to be less sensitive to testing strategy.

## Methods

### Data Sources

Alberta Health Services is the single health authority for Alberta and provides publicly-funded health care on behalf of the Ministry of Health. During the outbreak, both Alberta Health and Alberta Health Services have released regular online reports and conducted frequent press conferences.^7,8^ These reports included daily counts of new cases and cumulative numbers of laboratory tests performed (total tests and number of people tested), lab-confirmed cases, COVID-19-related hospital admissions and admission to intensive care units (ICU), and COVID-19-related deaths.^7,8^ Laboratory testing for SARS-CoV-2 virus has been based on a real-time reverse transcriptase–polymerase chain reaction (RT-PCR) assay using pharyngeal swab specimens. Details of the policy measures implemented by Alberta Health and Alberta Health Services are available online.^9^ We used additional sources to gather summary data on cumulative numbers of tests performed, and counts of laboratory-confirmed cases, recovered cases and deaths for all Canadian provinces.^3-6,10,11^

### Analysis

#### Overall approach

We used standard graphical methods to summarize data (bars, forest plots and map plots). Counts of lab-confirmed cases and hospitalization data were available across age groups and this information was used to assess potential associations between age and outcomes. We used packages developed in R statistical software to estimate R_0_ and transmissibility parameters.^15^

#### Modelling

We studied the basic reproductive number, R_0_, using information on *active cases* during the early phase of the outbreak before the declaration of the state of emergency in each province. R_0_ is the expected number of cases directly generated by one case in a population where all individuals are susceptible to infection. We estimated R_0_ using exponential growth and maximum likelihood methods.^14,16,17^ Since there are no primary data from Canada on the time interval between the onset of symptoms in the primary case and the onset of symptoms in a secondary case (serial interval, SI),^18^ we assumed a gamma distribution for SI, with a mean of 4.7 days and standard deviation of 2.9 days in main analysis based on Chinese data.^19^

We used log-linear models to estimate the parameters of the epidemic curves, i.e. rate of increase in positive cases per day and doubling-time in days. We planned to assess both the growth and decay phases of the outbreak, depending on its course at the time of this reporting.^20,21^

We considered two formal measures of transmissibility, the effective reproductive number (R_t_) and the infectiousness or force of infection (lambda). R_t_ is the average number of secondary cases that would be produced by a primary case infected at time t, if conditions remained constant after time t. R_t_ is useful to monitor the changes in the transmissibility of the epidemic over time in response to public health interventions.^22,23^ We used a Bayesian approach to estimate *R*_t_, given the time series of *daily incident*, *lab-confirmed cases* and the distribution of SI. Lambda estimates the daily effective infectiousness, subject to public health controls, with a projection of its decline if the outbreak is brought under control.^15,21^

#### Sensitivity analyses

To assess the effect of different assumptions for SI on the estimated value of the basic and effective reproductive numbers, we re-estimated R_0_ and R_t_ drawing the values of SI from gamma distributions with different parameters, which we obtained from the literature.^19, 24-26^ We also repeated the transmissibility analyses assuming that the observed rate of confirmed cases (ascertainment rate) was a variable fraction of the true COVID-19 cases in the province, i.e. ⅓, ½ and ⅔ during the first, second and third two-week period of the outbreak, respectively. In these analyses we used the five-day moving averages of new incident cases to minimize the noise generated by random daily fluctuations or backlogs of cases cleared at uneven intervals.

#### Hospitalization and death vs lab-confirmed cases

More aggressive policy measures and progressively broader indications for laboratory testing of symptomatic people may have influenced the detection of laboratory positive results in an unpredictable manner. For this reason, we considered both observed rates of laboratory positive cases, and hospitalization and death rates when interpreting the changes in measures of transmissibility and infectiousness during outbreak.

### Results

#### Course of the outbreak

As of May 7^th^, 255 people have been admitted to hospital and 114 have died in Alberta. Over 160,000 symptomatic people (3.7% of Albertans) were tested and over 6,000 had a positive result (0.14% of Albertans), with a peak of 319 new daily cases on April 23 (Figure 1 and eFigure 1). Most people who had lab-confirmed, positive tests were 30-49 years of age; the majority of those admitted to hospital were 50-years of age, or older (Figure 2). The Calgary Zone was the most affected health region in Alberta (eFigure 2). Alberta was the third province to confirm a case of COVID-19 and the third province to declare a state of emergency (Table 1). Policy interventions (eTable 1) were similar across the provinces and were all implemented rapidly, between March 12 and March 30 (eTable 2). Testing rates varied across provinces from 1.7% people tested in British Columbia to 3.8% in Alberta to (Table 1). Alberta had the third highest cumulative rate of positive cases, after Quebec and Ontario. Mortality rates in British Columbia and Alberta were similar and were lower than those in Ontario or Quebec (Table 2). Mortality rates increased with increasing rates of lab-confirmed cases in Ontario and Quebec, but not in Alberta (Figure 3).

**Figure 1:**
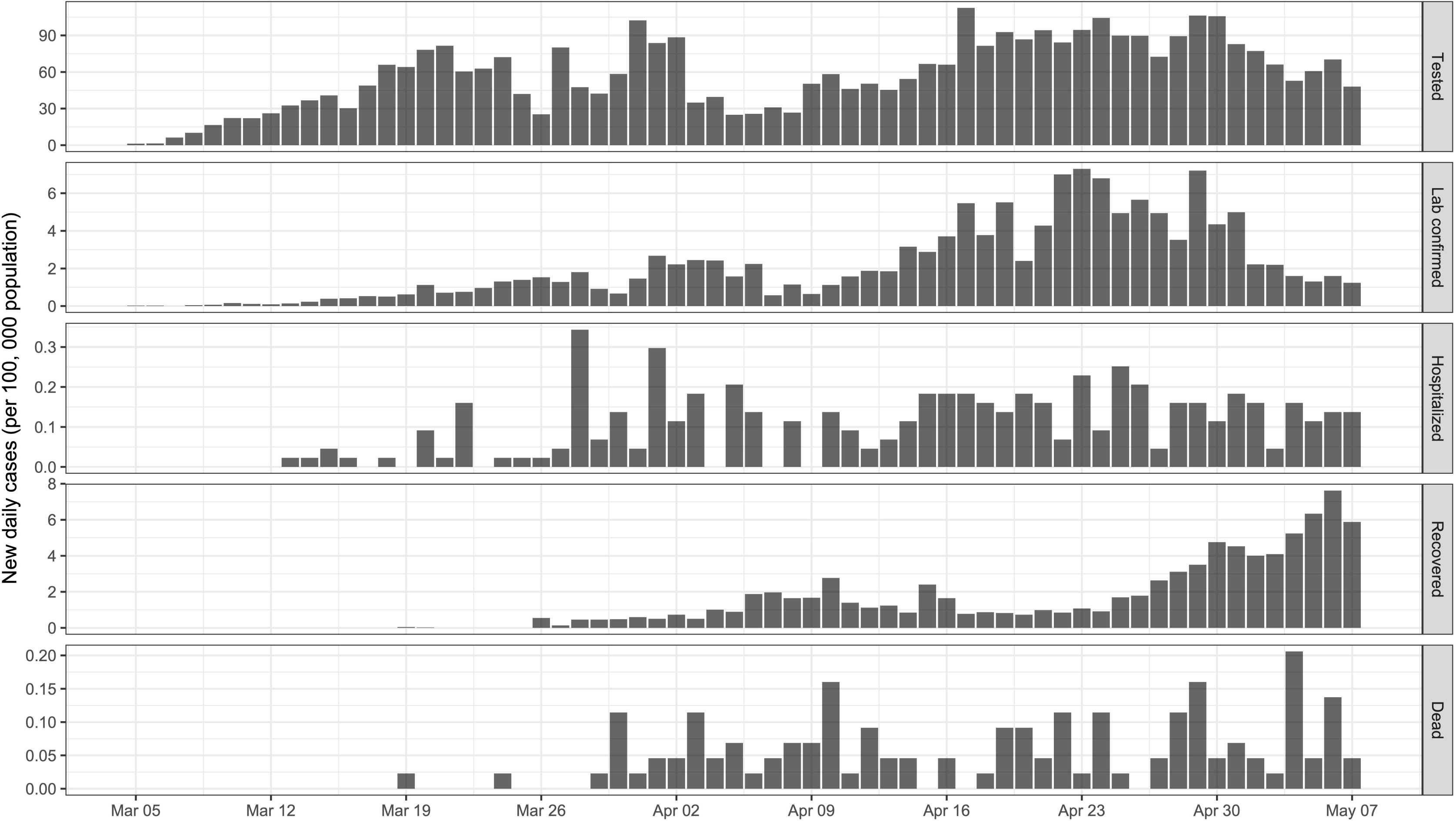
Incident cases. Legend: Rates of people tested (Tested), confirmed positive cases (Lab confirmed), COVID-19-related hospital admissions (Hospitalized), recovered from COVID-19 (Recovered) and COVID-19-related deaths (Dead). Rates are expressed as daily new cases per 100,000 population.

**Figure 2:**
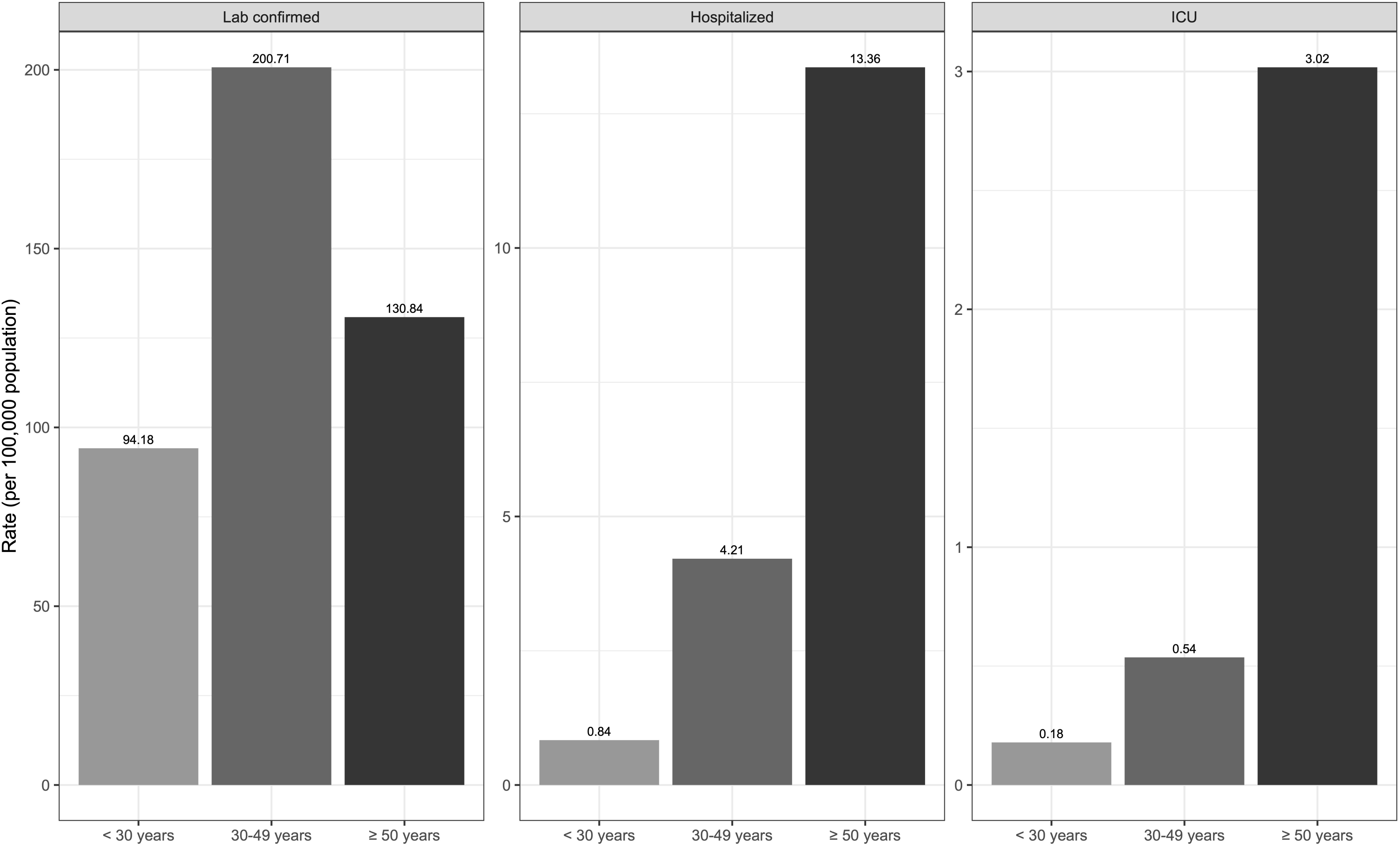
Incident rates in Alberta, British Columbia, Ontario and Quebec. Legend: Rates of confirmed positive cases (Lab confirmed) and COVID-19-related deaths (Dead). Rates are expressed as cumulative counts per 100,000 population.

**Figure 3:**
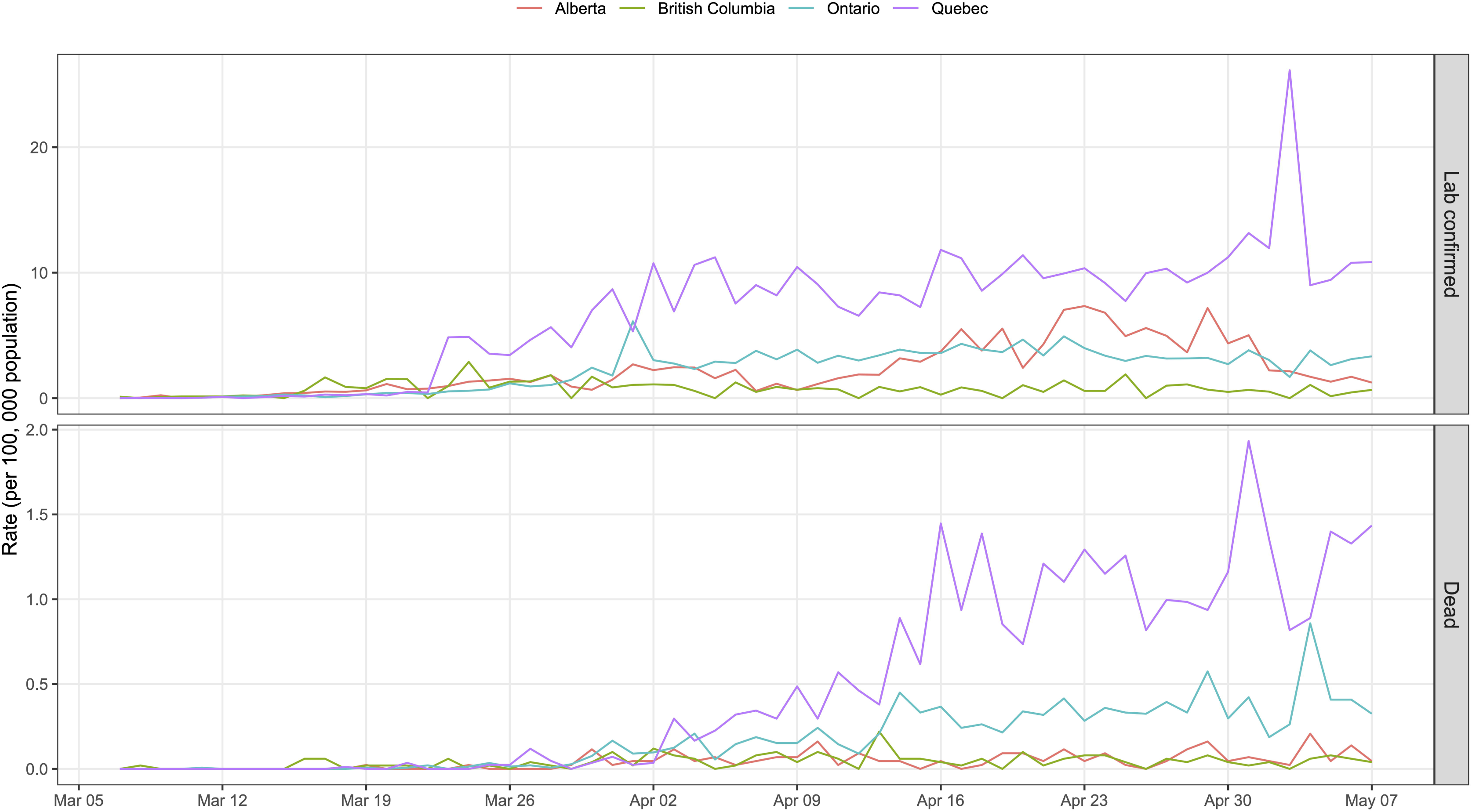
Outcomes by age. Legend: Rates of confirmed positive cases (Lab confirmed), COVID-19-related hospital admissions (Hospitalized) and COVID-19-related intensive care unit admissions (ICU). Rates are expressed as cumulative counts per 100,000 population.

**Table 1:** Index case, date of emergency declaration and lab tests by province as of May 7, 2020

| Province | Population | Date of the first case | Emergency declaration | Total tests (per 100,000 pop) | People tested (per 100,000 pop) |
| --- | --- | --- | --- | --- | --- |
| Alberta | 4,345,737 | March 5 | March 17 | 4,011 | 3,763 |
| British Columbia | 5,020,302 | January 28 | March 18 | 2,046 | 1,748 |
| Manitoba | 1,360,396 | March 12 | March 20 | 2,118 | 2,075 |
| Saskatchewan | 1,168,423 | March 12 | March 18 | 2,932 | 2,732 |
| Ontario | 14,446,515 | January 25 | March 17 | 2,636 | 2,550 |
| Quebec | 8,433,301 | February 27 | March 12 | 3,155 | 3,080 |
| New Brunswick | 772,094 | March 12 | March 19 | 2,153 | 2,033 |
| Newfoundland and Labrador | 523,790 | March 14 | March 18 | 1,806 | 1,804 |
| Nova Scotia | 965,382 | March 15 | March 22 | 3,527 | 3,474 |
| Prince Edward Island | 154,748 | March 14 | March 16 | 2,333 | 2,294 |

Sources: Berry et al.<sup>10</sup> and Government of Canada.<sup>11</sup>

**Table 2:**
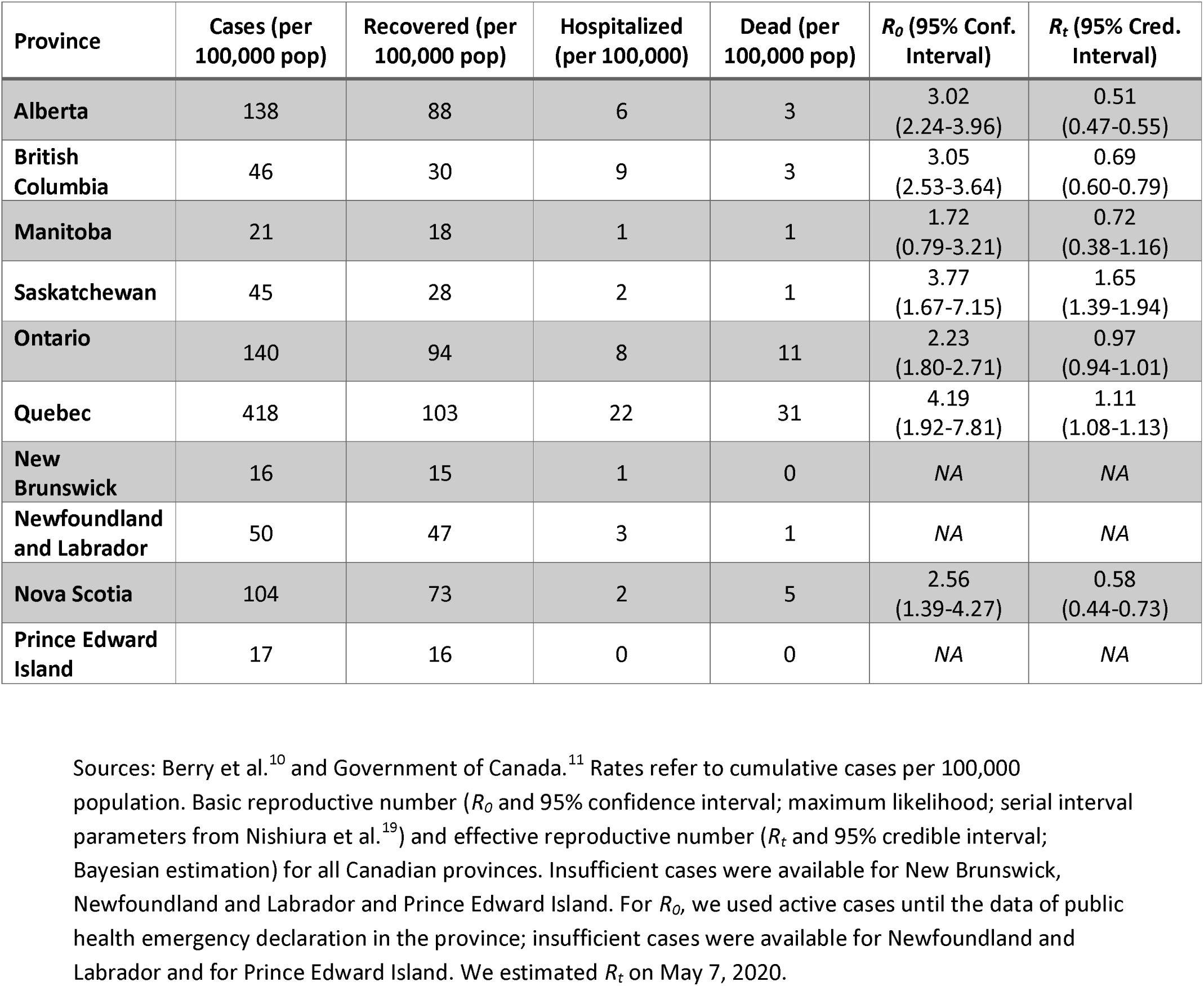
COVID pandemic data by province as of May 7, 2020

| Province | Cases (per 100,000 pop) | Recovered (per 100,000 pop) | Hospitalized (per 100,000) | Dead (per 100,000 pop) | $R_0$ (95% Conf. Interval) | $R_t$ (95% Cred. Interval) |
| --- | --- | --- | --- | --- | --- | --- |
| Alberta | 138 | 88 | 6 | 3 | 3.02<br>(2.24-3.96) | 0.51<br>(0.47-0.55) |
| British Columbia | 46 | 30 | 9 | 3 | 3.05<br>(2.53-3.64) | 0.69<br>(0.60-0.79) |
| Manitoba | 21 | 18 | 1 | 1 | 1.72<br>(0.79-3.21) | 0.72<br>(0.38-1.16) |
| Saskatchewan | 45 | 28 | 2 | 1 | 3.77<br>(1.67-7.15) | 1.65<br>(1.39-1.94) |
| Ontario | 140 | 94 | 8 | 11 | 2.23<br>(1.80-2.71) | 0.97<br>(0.94-1.01) |
| Quebec | 418 | 103 | 22 | 31 | 4.19<br>(1.92-7.81) | 1.11<br>(1.08-1.13) |
| New Brunswick | 16 | 15 | 1 | 0 | NA | NA |
| Newfoundland and Labrador | 50 | 47 | 3 | 1 | NA | NA |
| Nova Scotia | 104 | 73 | 2 | 5 | 2.56<br>(1.39-4.27) | 0.58<br>(0.44-0.73) |
| Prince Edward Island | 17 | 16 | 0 | 0 | NA | NA |

Sources: Berry et al.<sup>10</sup> and Government of Canada.<sup>11</sup> Rates refer to cumulative cases per 100,000 population. Basic reproductive number ( $R_0$ and 95% confidence interval; maximum likelihood; serial interval parameters from Nishiura et al.<sup>19</sup>) and effective reproductive number ( $R_t$ and 95% credible interval; Bayesian estimation) for all Canadian provinces. Insufficient cases were available for New Brunswick, Newfoundland and Labrador and Prince Edward Island. For $R_0$ , we used active cases until the data of public health emergency declaration in the province; insufficient cases were available for Newfoundland and Labrador and for Prince Edward Island. We estimated $R_t$ on May 7, 2020.

#### Impact of policy measures on the epidemic curve

Figure 4 summarizes the estimates of R_0_ during the early phase of the infection in Alberta (March 5-17, 2020). Estimates were similar by estimation method, but varied depending on the underlying distributional parameters for the SI from 2.6 (95% Confidence Interval 1.9-3.4) to 6.4 (4.3-8.5). Using the same estimation approach and assuming the same SI distribution, estimates of R_0_ varied across Canadian provinces, from 1.7 (0.78-3.2) for Manitoba to 4.2 (1.9, 7.8) for Quebec (Table 2). In Alberta, new cases increased at a rate of 9% per day (95% confidence interval 8-11%) before the peak. After the peak, new cases decreased at a rate of 8% per day (1-14%; eFigure 3).

**Figure 4:**
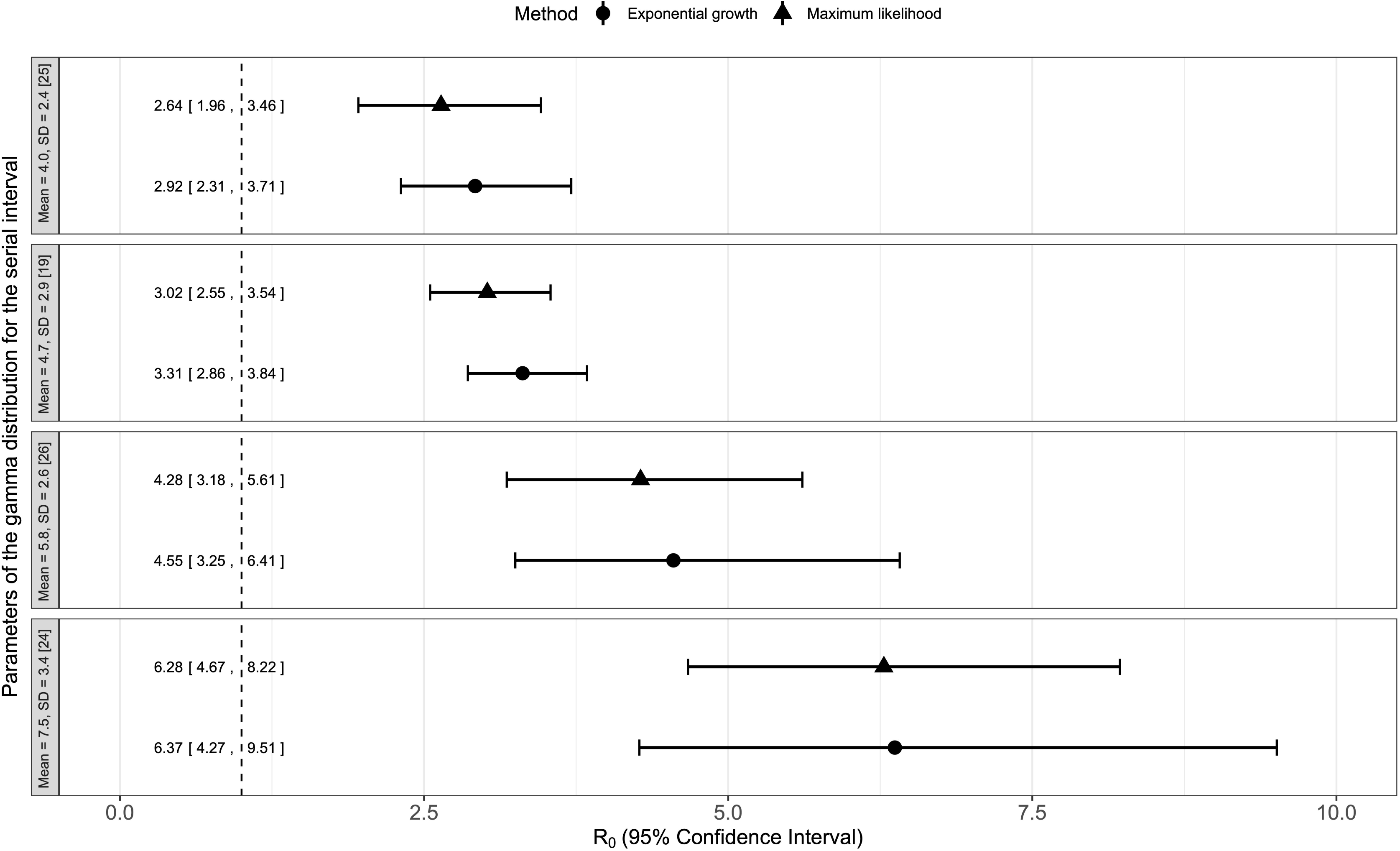
Basic reproductive number. Legend: The basic reproductive number (R_0_) was estimated between the first case and the date of emergency declaration (March 17, 2020). Estimates were obtained using two methods (maximum likelihood and exponential growth) and considering different parameters for the gamma distribution of the serial interval that were recently reported.^19,24-26^

#### Measure of transmissibility vs hospitalization and death

In Alberta, the curve of the effective reproduction number had an initial downward slope and a two-wave pattern. R_t_ was 2.67 (95% Credible Interval 2.13-3.27) on March 17 when the public health emergency was declared and declined sharply after public health measures intensified (Figure 5). R_t_ increased above 1 following the identification of localized outbreaks. These were tracked in three seniors homes, one oil sands plant, two meat-packing plants and one First Nations community, and accounted for 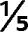 of the total provincial cases as of April 17.^7-8^ Accordingly, the global force of infection fell initially and subsequently rose (eFigure 4). During this later period of the outbreak, hospital admissions and mortality rates did not change. Measures of transmissibility did not change in sensitivity analyses (eFigures 4-5). Current data indicate that the outbreak is under control in Alberta, British Columbia and Nova Scotia (R_t_ <1; Table 2).

**Figure 5:**
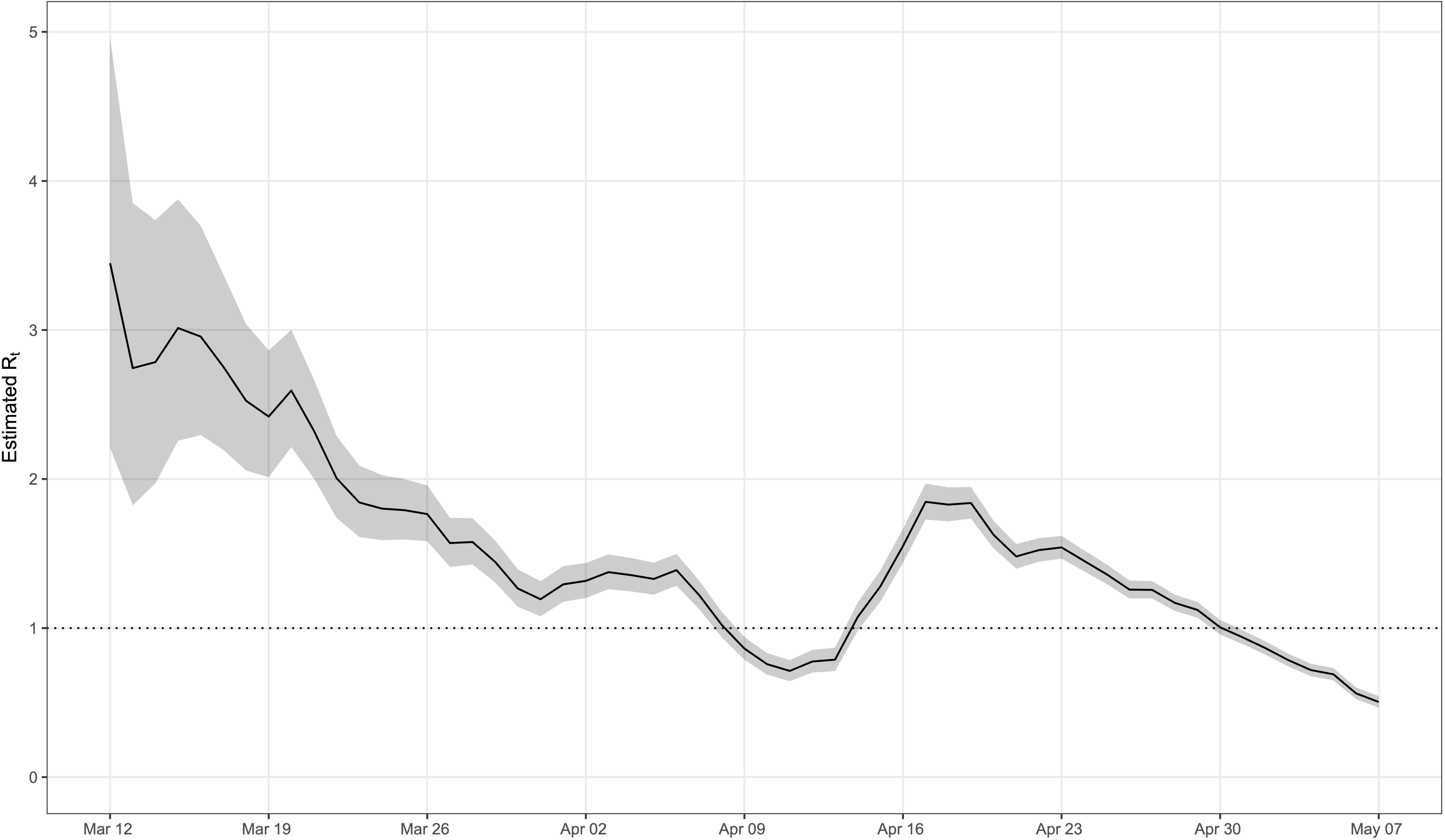
Effective reproductive number in Alberta. Legend: The effective reproduction number R_t_ is defined as the mean number of secondary cases generated by a typical primary case at time t in a population. The dotted horizontal line indicates R_t_ = 1, below which sustained transmission is unlikely if anti-transmission measures are sustained, indicating that the outbreak is under control. The 95% credible intervals are presented as gray shading. As of May 2, 2020, estimates of R_t_ were significantly <1.

## Discussion

Following the first confirmed COVID-19 case on March 5, the government of Alberta implemented aggressive measures to reduce interpersonal contacts, case importation and virus transmission in the population. All Canadian provinces rapidly implemented similar health-policy interventions, which successfully contained the outbreak. Measures of transmissibility dropped from the early phase of the pandemic to reproductive numbers close to or below 1. However, our study has identified many questions that need to be addressed if we want to be prepared for a second wave of COVID-19 or another pandemic in the future. First, initial estimates of the basic reproductive number were based on assumptions that may not hold. Estimates of the serial interval (time interval between the onset of symptoms in the primary case and the onset of symptoms in a secondary case) were obtained from world regions with different population density, culture and social structure. Canadian estimates are needed to obtain more accurate estimates of viral transmissibility and plan actions during the early phase of an outbreak. Second, hospitalization and mortality outcomes varied by province for reasons that were potentially unrelated to the health-policy measures implemented. Studies are needed to investigate how the public receive and respond to these interventions. Third, without clear and consistent testing criteria, it is difficult to rely on rates of positive cases to predict health outcomes. More information on current testing strategy should become available during the course of an outbreak.

The findings of this study are reassuring as efforts to combat the global pandemic of COVID-19 seem to successfully control the outbreak in Canada,^5^ similar to what was reported in China.^22,27^ Vigorous and multifaceted measures of containment, mitigation and suppression along with extensive public education are associated with improved control of the epidemic in the absence of an effective pharmacological intervention. These measures included traffic minimization, cancellation of social gatherings, home quarantine, and reorganization of health care services. Although these interventions rapidly prevented the surge of severe cases requiring hospital admissions in most provinces, local spread has been contained only recently and vigilance needs to remain in place to avoid under-ascertainment and/or under-reporting and to ensure existing outbreaks of COVID-19 are brought under full control.

Our data do not support the association between early arrival of the pandemic, complexity of organizations administering health care or low testing rates with poor outbreak outcomes. Although the pandemic arrived in Alberta approximately two months after the first outbreak in China, providing the government and the public the advantage of preparing a response,^1,3^ the outbreak had a different course in British Columbia and Ontario, where the first case was identified around the same time. Also, the course of the outbreak was similar in provinces with a single health care body (Alberta) and multiple organizations (British Columbia), suggesting that policy responses can be efficiently coordinated and delivered regardless of the system complexity. Finally, although Alberta data are consistent with those of countries that performed massive, population-wide testing and adopted more aggressive policies,^28-36^ British Columbia had the lowest testing rate in Canada and successfully contained the outbreak. In addition to prompt prevention and management strategies, as well as high quality and coordinated information delivery and public education and response, other factors may have played a role. Of the three major lineages of SARS-COV-2 a recent phylogenetic genome network analysis identified,^37^ the virus type found in Canada is the same that emerged from bats and pangolins and originally affected Wuhan, which may be more benign than the types predominant later in Wuhan and China, in the USA or Europe. While a more benign viral type may have been responsible for the Canadian outbreak, further studies are needed to validate this claim.

Our data on the association between age and poorer clinical outcomes are consistent with studies showing that COVID-19 is more lethal in older patients and people with underlying comorbid conditions.^12^ Disease transmissibility has been extensively investigated. Studies have reported values of R_0_ ranging from 1.4 to 6.5, depending on the data sources, time periods and model assumptions. ^24 27, 28^ The overall, downward slope of the effective reproduction number curve suggests that containment efforts were succeeding in progressively reducing transmission of the infection in Alberta. Role of asymptomatic people in the infection spread at the beginning of the outbreak, uncertain serial intervals or incubation periods may explain imprecise initial estimates of disease transmissibility. The wide range of the estimates of the basic reproductive number across Canadian provinces are consistent with this finding. The two-wave pattern we observed in Alberta reflects the occurrence of localized outbreaks, which imply that continued vigilance is needed. Individual-level protection strategies for vulnerable people including residents of long-term care facilities, and for heavily-crowded industrial areas are needed to prevent second outbreaks in Alberta, British Columbia and Nova Scotia, and bring the outbreak in the remaining provinces under control, where the reproductive number is not <1 yet. These considerations are of paramount importance for phased relaunching strategies and easing of restrictions currently being planned in some provinces.^7^

Our study has several limitations. First, individual strategies could not be evaluated as all provincial governments implemented multiple interventions early, and over a short time-period, to control the outbreak. Second, as in any observational design, causal inference cannot be made, and the relationship between intervention and outcomes can only be considered associative. Third, limited information was available about incubation period, serial interval, time to key outcomes and clinical characteristics of people who developed COVID-19. Finally, we had no information to minimize possible ascertainment bias and the rate of asymptomatic cases, and hospitalization data were not available for all provinces. While we believe that these limitations do not affect our results and interpretations, we do believe data collection and access are areas where new strategies are needed to improve the management of public health emergencies.

In summary, the implementation of multifaceted health policy interventions was associated with a relatively rapid control of the COVID-19 outbreak in Alberta, British Columbia and Nova Scotia. The outbreak is not yet under control in other provinces. Our study shows that depending on the virulence of a virus or the effectiveness of the health policy interventions, hard outcome measures, including hospitalization and intensive care admission may be a more reliable indicator of how successfully an outbreak is controlled than rates of positive test results.

## Funding statement / Competing interest

none (details at the end of the manuscript)

## Data Availability

Data used a publicly available

## Contributions

Study concept and design, analysis plan: Alexis Guigue, David Vickers, MD Mahsion, Pietro Ravani, Seungwon Lee

Data collection: Alexis Guigue, Seungwon Lee

Analysis: MD Mahsin, Pietro Ravani, David Vickers

Manuscript draft: MD Mahsin, Pietro Ravani, Seungwon Lee

Manuscript revision: all authors

Contributed to intellectual content: all authors

Study supervision: Pietro Ravani

## Transparency

All authors affirm that this manuscript is an honest, accurate, and transparent account of the study being reported; that no important aspects of the study have been omitted; and that any discrepancies from the study as planned have been explained.

## Funding

This study was not supported by any specific funding.

## Competing interests

All authors declare: no support from any organization for the submitted work; no financial relationships with any organizations that might have an interest in the submitted work in the previous three years; no other relationships or activities that could appear to have influenced the submitted work.

## Ethics approval

Not required.

## Data sharing

We used publicly available data and referred to the sources that are publicly available.

## Disclaimer

The interpretation and conclusions contained herein are those of the researchers and do not represent the views of the Government of Alberta, Alberta Health, Alberta Health Services, the Cumming School of Medicine or the University of Calgary.

## Exclusive license

The Corresponding Author has the right to grant on behalf of all authors and does grant on behalf of all authors, a worldwide licence to the CMAJ.

## Notes

### Competing Interest Statement

The authors have declared no competing interest.

